# Plasma extracellular vesicle transcriptome as a dynamic liquid biopsy in acute heart failure

**DOI:** 10.1101/2023.02.17.23285936

**Authors:** Priyanka Gokulnath, Michail Spanos, H. Immo Lehmann, Quanhu Sheng, Rodosthenis Rodosthenous, Mark Chaffin, Dimitrios Varrias, Emeli Chatterjee, Elizabeth Hutchins, Guoping Li, George Daaboul, Farhan Rana, Ashley Mingyi Wang, Kendall Van Keuren-Jensen, Patrick T. Ellinor, Ravi Shah, Saumya Das

## Abstract

**Background:** Acute decompensation is associated with increased mortality in heart failure (HF) patients, though the underlying etiology remains unclear. Extracellular vesicles (EVs) and their cargo may mark specific cardiovascular physiologic states. We hypothesized that EV transcriptomic cargo, including long non-coding RNAs (lncRNAs) and mRNAs, is dynamic from the decompensated to recompensated HF state, reflecting molecular pathways relevant to adverse remodeling.

**Methods:** We examined differential RNA expression from circulating plasma extracellular RNA in acute HF patients at hospital admission and discharge alongside healthy controls. We leveraged different exRNA carrier isolation methods, publicly available tissue banks, and single nuclear deconvolution of human cardiac tissue to identify cell and compartment specificity of the topmost significantly differentially expressed targets. EV-derived transcript fragments were prioritized by fold change (−1.5 to + 1.5) and significance (<5% false discovery rate), and their expression in EVs was subsequently validated in 182 additional patients (24 control; 86 HFpEF; 72 HFrEF) by qRT-PCR. We finally examined the regulation of EV-derived lncRNA transcripts in human cardiac cellular stress models.

**Results:** We identified 138 lncRNAs and 147 mRNAs (present mostly as fragments in EVs) differentially expressed between HF and control. Differentially expressed transcripts between HFrEF vs. control were primarily cardiomyocyte derived, while those between HFpEF vs. control originated from multiple organs and different (non-cardiomyocyte) cell types within the myocardium. We validated 5 lncRNAs and 6 mRNAs to differentiate between HF and control. Of those, 4 lncRNAs (AC092656.1, lnc-CALML5-7, LINC00989, RMRP) were altered by decongestion, with their levels independent of weight changes during hospitalization. Further, these 4 lncRNAs dynamically responded to stress in cardiomyocytes and pericytes *in vitro*, with a directionality mirroring the acute congested state.

**Conclusion:** Circulating EV transcriptome is significantly altered during acute HF, with distinct cell and organ specificity in HFpEF vs. HFrEF consistent with a multi-organ vs. cardiac origin, respectively. Plasma EV-derived lncRNA fragments were more dynamically regulated with acute HF therapy independent of weight change (relative to mRNAs). This dynamicity was further demonstrated with cellular stress *in vitro*. Prioritizing transcriptional changes in plasma circulating EVs with HF therapy may be a fruitful approach to HF subtype-specific mechanistic discovery.

**CLINICAL PERSPECTIVE:** *What is new?:* We performed extracellular transcriptomic analysis on the plasma of patients with acute decompensated heart failure (HFrEF and HFpEF) before and after decongestive efforts. Long non-coding RNAs (lncRNAs) within extracellular vesicles (EVs) changed dynamically upon decongestion in concordance with changes within human iPSC-derived cardiomyocytes under stress. In acute decompensated HFrEF, EV RNAs are mainly derived from cardiomyocytes, whereas in HFpEF, EV RNAs appear to have broader, non-cardiomyocyte origins.

*What are the clinical implications?:* Given their concordance between human expression profiles and dynamic *in vitro* responses, lncRNAs within EVs during acute HF may provide insight into potential therapeutic targets and mechanistically relevant pathways. These findings provide a “liquid biopsy” support for the burgeoning concept of HFpEF as a systemic disorder extending beyond the heart, as opposed to a more cardiac-focused physiology in HFrEF.

## INTRODUCTION

Despite consistent advances in chronic heart failure (HF) management with guideline-directed medical therapy, each admission for acute decompensated HF (ADHF) is accompanied by accelerated disease progression, mortality, debility, or need for advanced therapy.^1^ The mechanism of this clinical observation remains elusive, ranging from activation of systemic inflammatory and fibrotic pathways (affecting both cardiac and non-cardiac organs) during ADHF, inadequate decongestion, subclinical ischemia, and increased susceptibility to arrhythmia.^2^ In addition, this phenomenon is observed in both HF with “preserved” left ventricular ejection fraction (HFpEF, LVEF >/= 0.50) and HF with “mid-range” or “reduced” LVEF (HFrEF; LVEF </= 0.50),^3^ potentially implicating some systemic factor not directly related to cardiac function in its etiology. Indeed, broad quantification of the circulating proteome in patients with HF suggests a prominent role for inflammatory and immune activation in prognosis, comorbidity, and cardiac remodeling.^4^ Nevertheless, most biomarker efforts (outside of natriuretic peptides) have generally focused on <u>chronic</u> HF. Identification of molecules dynamically altered during ADHF with a potential to impact multi-organ function may provide important disease trajectory biomarkers and illuminate novel targetable pathways responsible for downstream prognostic implications of HF hospitalization.

While most “omics” studies of HF have relied on bulk plasma for discovery, a key limitation to these approaches is the absence of insight into tissue specificity and issues with reverse causation (whether the molecules are a cause or consequence of acute HF). Recent studies report that human genetic variation may not explain mechanistic differences across subtypes of HF (HFpEF vs. HFrEF).^5^ In addition, direct biopsy studies are usually conducted in a compensated or end-stage condition,^6,7^ limiting the discovery of modifiable disease mechanisms. Extracellular vesicles (EVs) are nano-sized lipid bilayered that carry various molecular cargo (proteins, DNA, RNA) that may both function “at a distance” in intercellular communication,^8–10^ and serve as potential biomarkers.^10,11^

Most prior studies have examined the small RNA content of EVs, given their presence in biofluids as full-length transcripts and their potential to mediate downstream regulation of post-transcriptional regulation in recipient cells.^10,12^ In addition to their potential functional role in signaling, the RNA content of EVs may mirror signaling in their tissue of origin,^10^ allowing EV transcriptomics to serve as a “liquid biopsy.” Indeed, mRNAs and long noncoding RNAs (lncRNAs) demonstrate more tissue-specific expression than small RNAs,^13–15^ but have not been investigated in detail. While EV RNA cargo may serve as a biomarker for ventricular remodeling in chronic HF state,^16^ these studies are limited by a single time point measure without large physiologic perturbation seen in the acute phase, thereby limiting detection of HF-related effects on the transcriptome. We have previously demonstrated the feasibility of measuring long RNA transcripts in plasma using RNA sequencing.^17^ Here, we characterize EV RNA cargo during ADHF hospitalization, demonstrating broad transcriptional differences across HFpEF and HFrEF and dynamic transcriptional changes in EV cargo during decongestion, independent of clinical-biochemical indices of congestion. These dynamic changes in candidate EV RNA cargo during HF therapy were validated in 182 patients. We used publicly available curated expression atlases, including single nuclear RNA sequencing in human cardiomyopathies, to deconvolute the cell type of origin in HFpEF-vs. HFrEF-derived EV transcripts during acute HF. To implicate the RNA cargo as a response to the HF decompensation in cardiomyocytes, we used human iPSC-derived cellular models under stress that demonstrate dynamic alterations in EV and cellular lncRNA expression. Ultimately, these investigations aimed to characterize the profile, plasticity, and etiology of EV transcriptional differences during ADHF across HF sub-types, as a novel “liquid biopsy” for the cardiovascular state that may prioritize future discovery.

## METHODS

### Study cohort

Between September 2016 and May 2019, 181 patients with ADHF (discovery, N=14; validation, N=167). ADHF diagnosis required (1) clinical symptoms (including exertional or rest dyspnea, orthopnea, or PND) and biochemical evidence (N-terminal pro-BNP level > 1000 pg/ml or BNP > 400 pg/ml), or (2) clinical signs: radiographic pulmonary edema or pleural effusions, elevated jugular venous pressure, lower extremity edema, or rales on pulmonary examination, hemodynamic evidence (right atrial pressure > 10 mmHg; pulmonary capillary wedge pressure > 18 mmHg) and clinical response to intravenous diuretic therapy (as determined by a physician). Additional inclusion criteria were age >/= 18 years and an assessment of left ventricular function within the last year or planned during hospital admission. LVEF was acquired non-invasively within one day from admission, based on echocardiography readings. LVEF was acquired by end-diastolic frame minus net counts in the end-systolic frame divided by net counts in end-diastole and confirmed by another clinician using recalculation in 2D mode (DV). Patients with cardiac amyloidosis or active malignancies were excluded. Diuretic therapy was determined by treating providers. As a healthy “control” sample, we included 33 patients who underwent ablation for supraventricular tachycardia (SVT), with blood available at the time of ablation (discovery, N=9; validation, N=24; **Table 1 and 2**) with normal cardiac function by clinical echocardiographic study in the past year. The study protocol was approved by the Institutional Review Board at Massachusetts General Brigham.

**Table 1.**
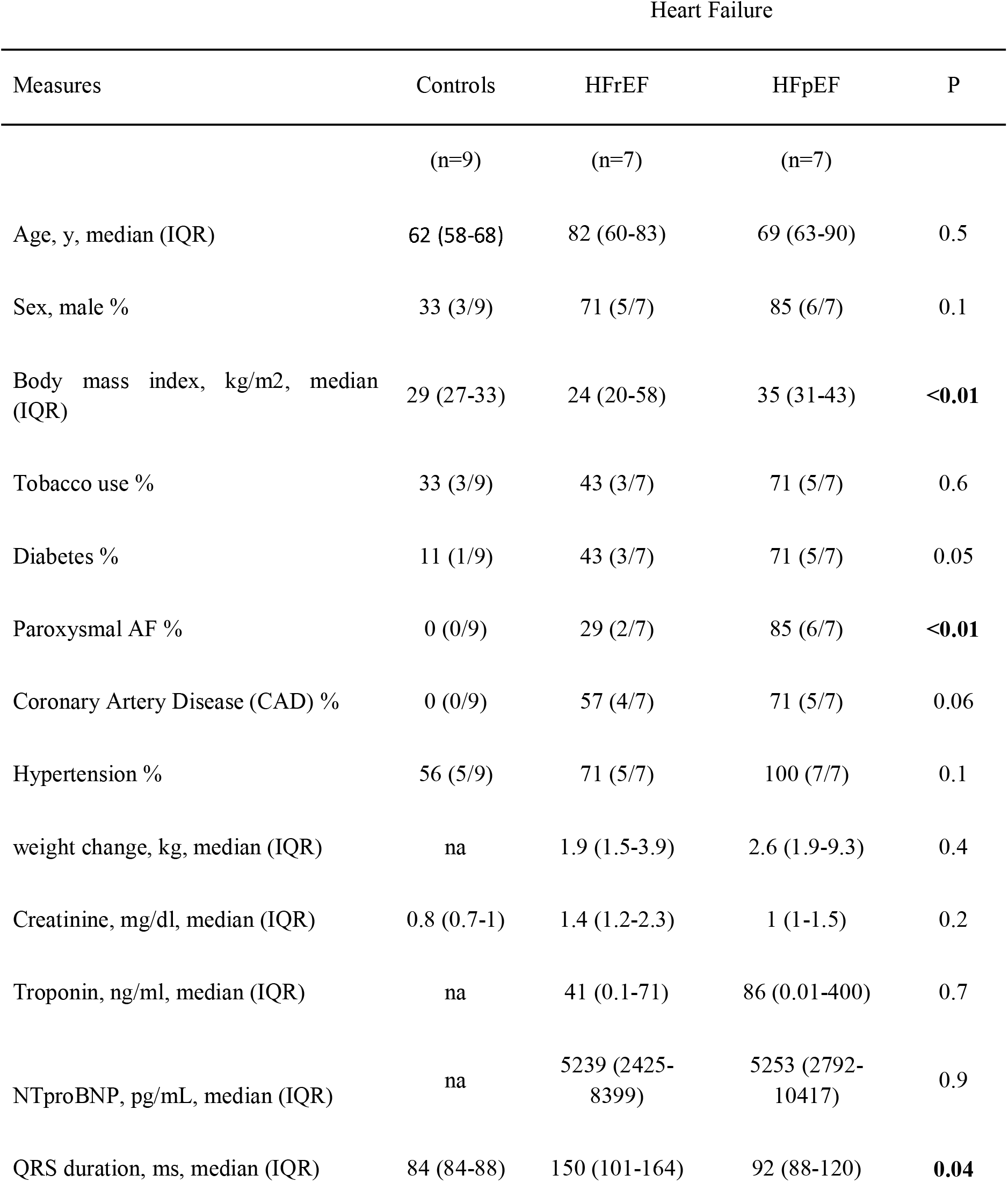
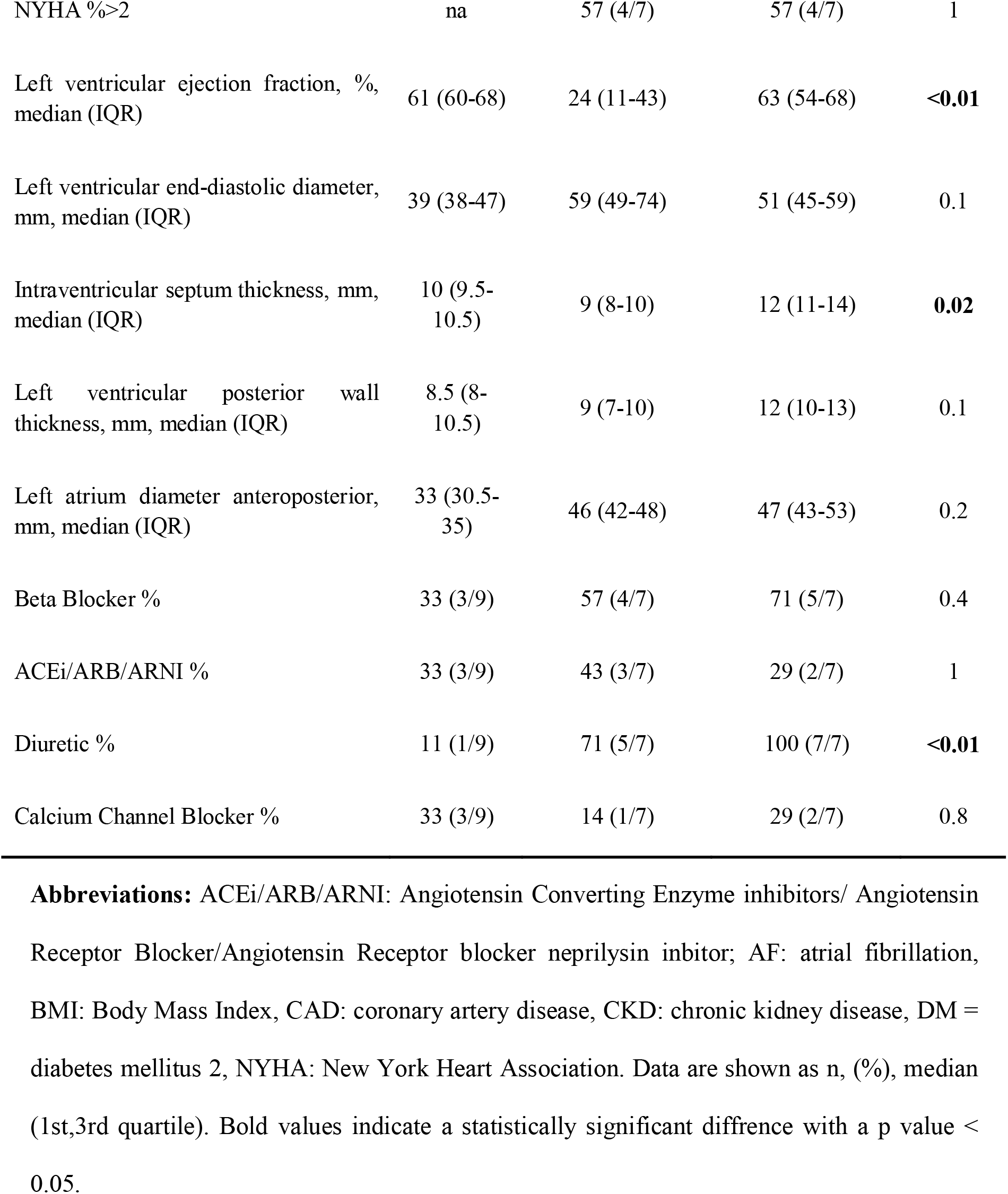
Baseline Characteristics of the discovery cohort.

**Table 2.**
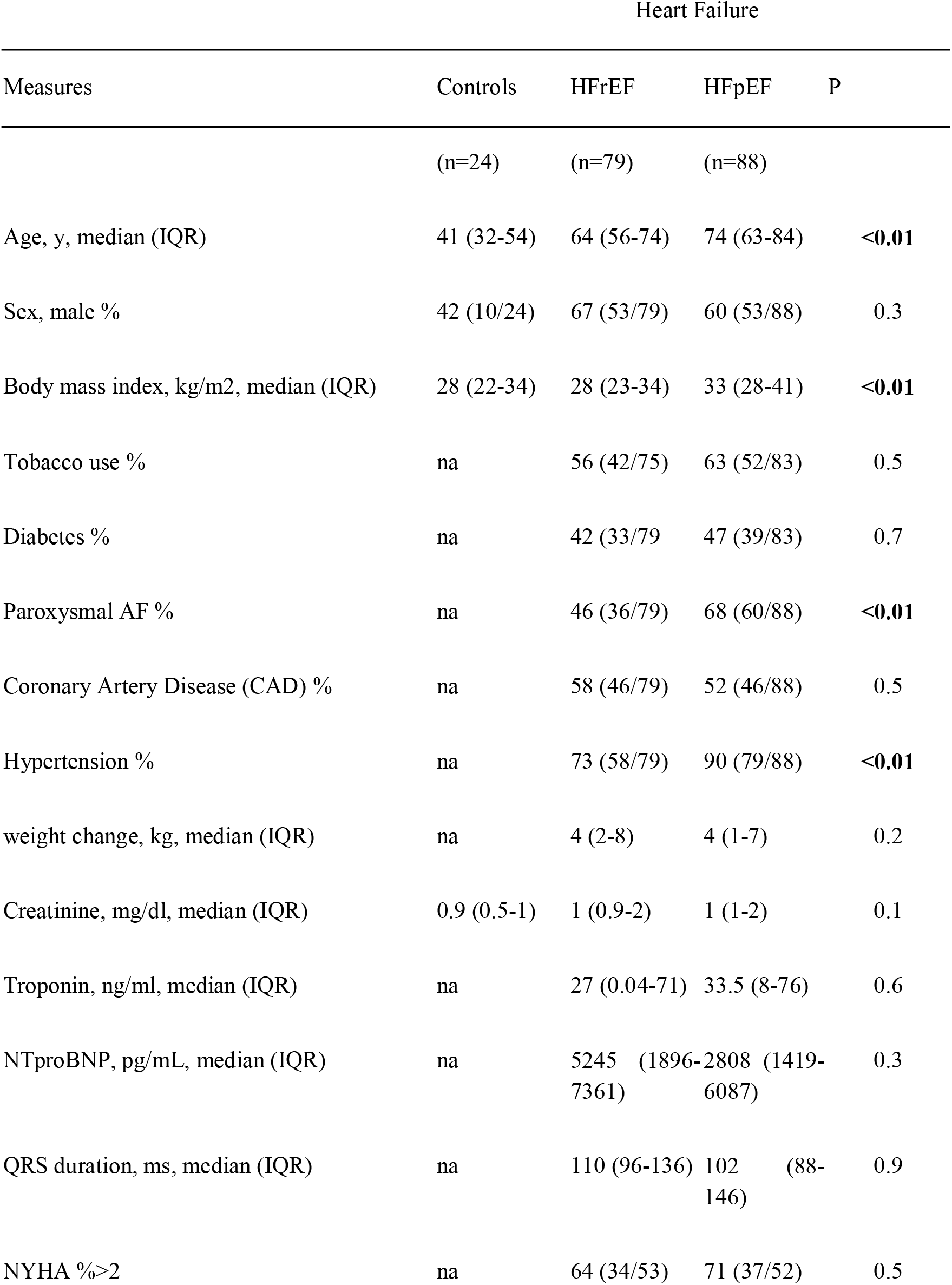
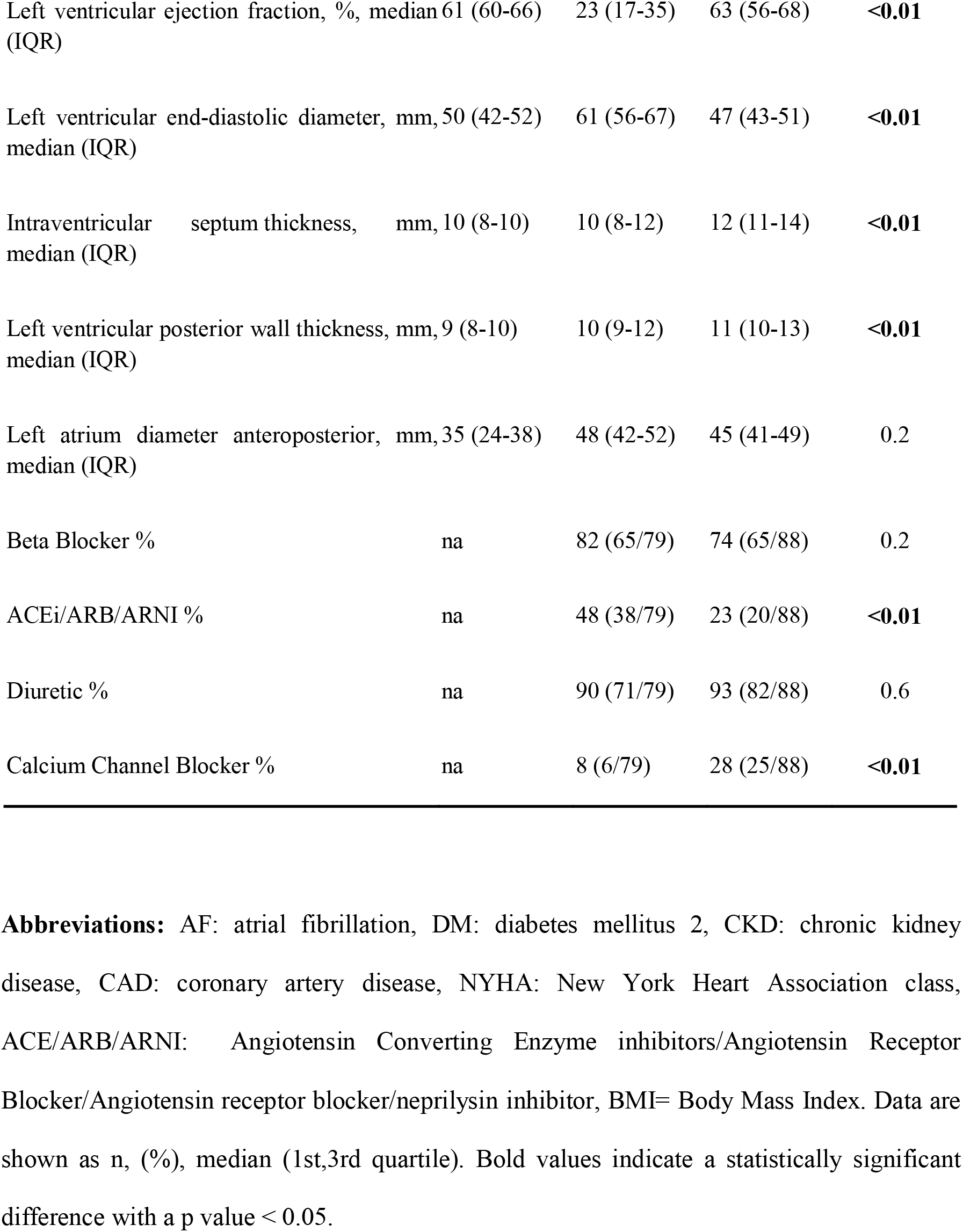
Baseline Characteristics of the validation cohort.

### Extracellular RNA isolation and sequencing

Extracellular RNA (ExRNA) was isolated from the plasma of 37 patients in our discovery cohort (Control, n=9; HFrEF, n=7 at admission and discharge; HFpEF, n=7 at admission and discharge) using the miRNeasy Serum/Plasma kit (QIAGEN, Germantown, MD, USA) as per manufacturer’s protocol, cDNA libraries were constructed using the SMARTer Stranded Total RNA-Seq Kit v2 Pico Input Mammalian (Takara Bio, San Jose, CA, USA) and sequenced using NextSeq 2000 platform (Illumina, San Diego, CA, USA). Analysis was performed on methods detailed in the Supplementary material. All the significant genes are compiled in **Supplementary Table1**. Tissue-enrichment analysis was performed using an in-house R script with tissue-wise RNA expression data available from the Human Protein Atlas and using TissueEnricher webtool.

### RNA isolation from different plasma compartments

RNA was isolated from each of the following compartments previously noted to be the main carriers of RNA in biofluids^18^ (EV compartments, lipid fractions, AGO fraction) from a starting volume of 0.5 mL pooled plasma (control and heart failure samples) for each replicate using the detailed protocol is provided in the Supplementary material. Each fraction was validated for its characteristic markers using a western blot (**Supplementary Figure 1**). EV compartment was validated using markers for Alix, CD63, CD81, Syntenin, 58K Golgi protein; lipid fractions by Apo A1 and Apo E, Ago fractions with Ago antibody. EV compartment characterization has been included in the supplementary material. cDNA synthesis, qRT-PCR and digital PCR methods are described in the Supplementary methods.

### Single nuclear RNA sequencing

To investigate cardiac cell type of origin and expression patterns, the expression of single cardiac nuclear transcriptome from 11 human hearts with dilated cardiomyopathy (DCM), 15 human hearts with end-stage hypertrophic cardiomyopathy (HCM), and 16 non-failing human hearts were analyzed^19^. The snRNAseq data was normalized and scaled. The topmost differentially expressed genes in plasma from HFpEF vs. Control EVs and HFrEF vs. Control EVs were filtered based on FDR (<5%) were analyzed using the scanpy function (scanpy.tl.score_gene) and then enriched nuclei were overlayed on a Uniform manifold approximation and projection (UMAP) plot from normal heart, DCM and HCM. These genes were also represented by a dot-plot of the different cardiac cell types generated using an in-house R script.

### *In vitro* (cellular) studies

Human induced pluripotent stem cell-derived cardiomyocytes (iPSC-CMs) were cultured according to the protocol mentioned in the study by Lian et al.^20^ Human cardiac fibroblasts (6330, ScienceCell, Carlsbad, CA, USA) and pericytes (C-12980, PromoCell, Heidelberg, Germany) were commercially obtained and cultured according to the manufacturer’s instructions. Cell stressors were performed as mentioned in the publication by Li et al.^21^ Hypoxia and nutrient deprivation (glucose serum deprivation) were each subjected for 5 hours in iPSC-CMs and 24 hrs for fibroblasts and pericytes. For EV RNA isolation, condition media was collected after the stress treatment, concentrated, and EV was isolated using the ExoRNAeasy kit. Cellular RNA was collected from cells using the Trizol method previously mentioned. EV and cellular RNA collected were further proceeded with cDNA synthesis and qRT-PCR as mentioned above.

### Statistical analysis

Statistical analyses of qPCR and dPCR were performed using GraphPad Prism software (Version 9, San Diego, CA, USA). qPCR data are expressed as mean ± SEM, and the statistical significance was assessed by Kruskal-Wallis test. All other statistical analysis was performed using R and further described in the supplementary material.

## RESULTS

### Cohort characteristics

Baseline clinical characteristics for our analytic cohorts are shown in **Tables 1** and **2**. The median (IQR) time between admission and initial blood draw was 2 (1-3) days for the discovery cohort and 3 (2-4) days for the validation cohort. The median (IQR) hospital stay was 6 (3-8) days in the discovery cohort and 5 (3-7) days in the validation cohort. The median (IQR) time between the final blood draw and discharge was 1 (1-3) day in both cohorts. In general, individuals with HFpEF had increased body mass index (median BMI [IQR], discovery cohort HFpEF 35 kg/m^2^, [35-43 kg/m^2^]; p<0.01), with a higher prevalence of atrial fibrillation (p<0.01). HFpEF patients in our validation cohort were older and had a higher prevalence of hypertension. Sixty-seven percent of HFrEF patients were male, 58% had ischemic cardiac disease and a history of CAD, while 38% had undergone coronary artery bypass graft surgery at the time of admission. Regarding volume status, HFpEF and HFrEF patients both reduced body weight following decongestion. As expected, the level of NT-proBNP was high in all patients during admission; however, decongestive treatment resulted in a significant decline in NT-proBNP levels only in HFrEF patients.

### Plasma transcriptional profiling in ADHF reveals broad differences across HF subtypes

The overall analytical flow is shown in **Figure 1**. We first performed long RNA sequencing of plasma extracellular RNA on our discovery cohort, as described in the methods section using the SMARTer Stranded Total RNA-Seq library kit. The use of this protocol allows for the discovery of both mRNA and lncRNA transcripts.^17^

**Figure 1.**
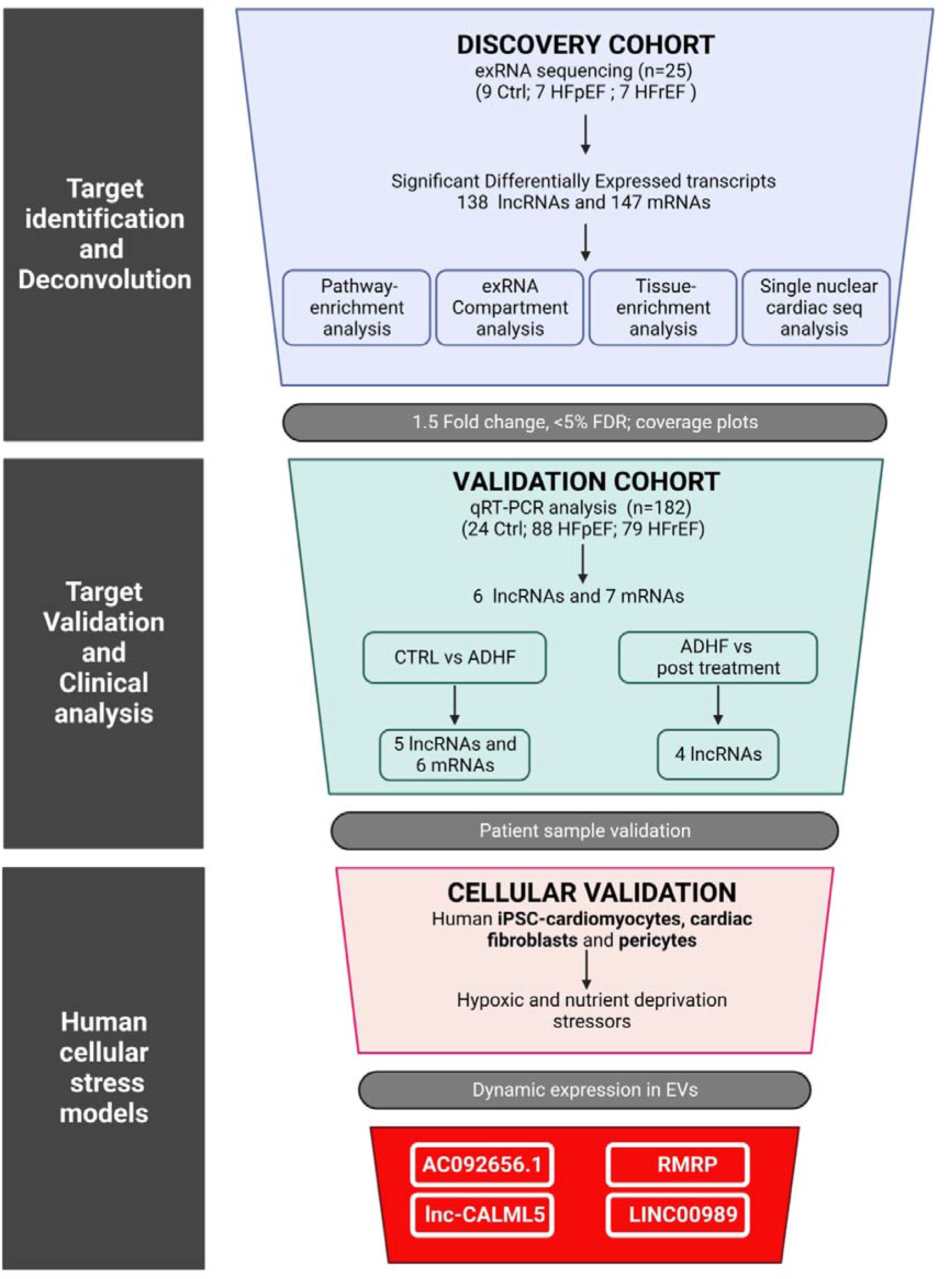
Flowchart of the study overview.

Of the total reads, an average of 53.4% was uniquely mapped to the human genome. Multi-mapped reads were omitted for the gene counting. On average, 10.4% of uniquely mapped reads were assigned to genes (**Supplementary table 1**). An average number of 14111 and 16697 mRNA transcripts and 13766 and 15580 lncRNA transcripts were identified (transcripts per million, TPM >= 1.0) in control and heart failure patient plasma samples, respectively. An average of 17045 and 16350 mRNA transcripts and 16590 and 14570 lncRNA transcripts were detected at ADHF state (Visit 1 or V1) and after decongestion therapy (V2), respectively (**Supplementary Table 3, Supplementary Figure 3**).

In unsupervised clustering of the transcripts profiled in ADHF (corresponding to V1) (**Figure 2A**), control and HF subtypes were grouped separately, with the long non-coding RNA transcriptome demonstrating more consistent differences between the groups. We observed broad transcriptional differences between ADHF states (both HFpEF and HFrEF) versus control patients (**Figure 2B**). Notably, we observed much broader differences in the transcriptome of HFpEF (compared to HFrEF), with a prominent contribution of lncRNAs. A total of 399 RNAs were increased in HFpEF plasma relative to control (208 mRNAs; 191 lncRNAs) and 100 transcripts decreased (74 mRNAs; 26 lncRNAs). In comparison, of 31 RNAs increased in HFrEF patient plasma relative to controls, 22 were mRNAs, and 9 were lncRNAs, while 7 transcripts were decreased (3 mRNAs and 4 lncRNAs) (**Figure 2C**). Strikingly, the differential transcriptome during congested ADHF was largely distinct between HFpEF and HFrEF compared to controls (**Figure 2D**), with most transcripts specific to a given HF subtype. Kyoto Encyclopedia of Genes and Genomes (KEGG) Pathway enrichment analysis showed that the differentially expressed transcripts in HFrEF were primarily associated with the heart, such as Dilated cardiomyopathy, Hypertrophic cardiomyopathy, adrenergic signaling in cardiomyocytes, etc. By contrast, in HFpEF, implicated pathways were variable and included a broad range of cellular signaling pathways, such as melanogenesis, Rap1 signaling pathway, chemokine signaling pathway, focal adhesion, and pathways involving cancer (**Figure 2E)**.

**Figure 2.**
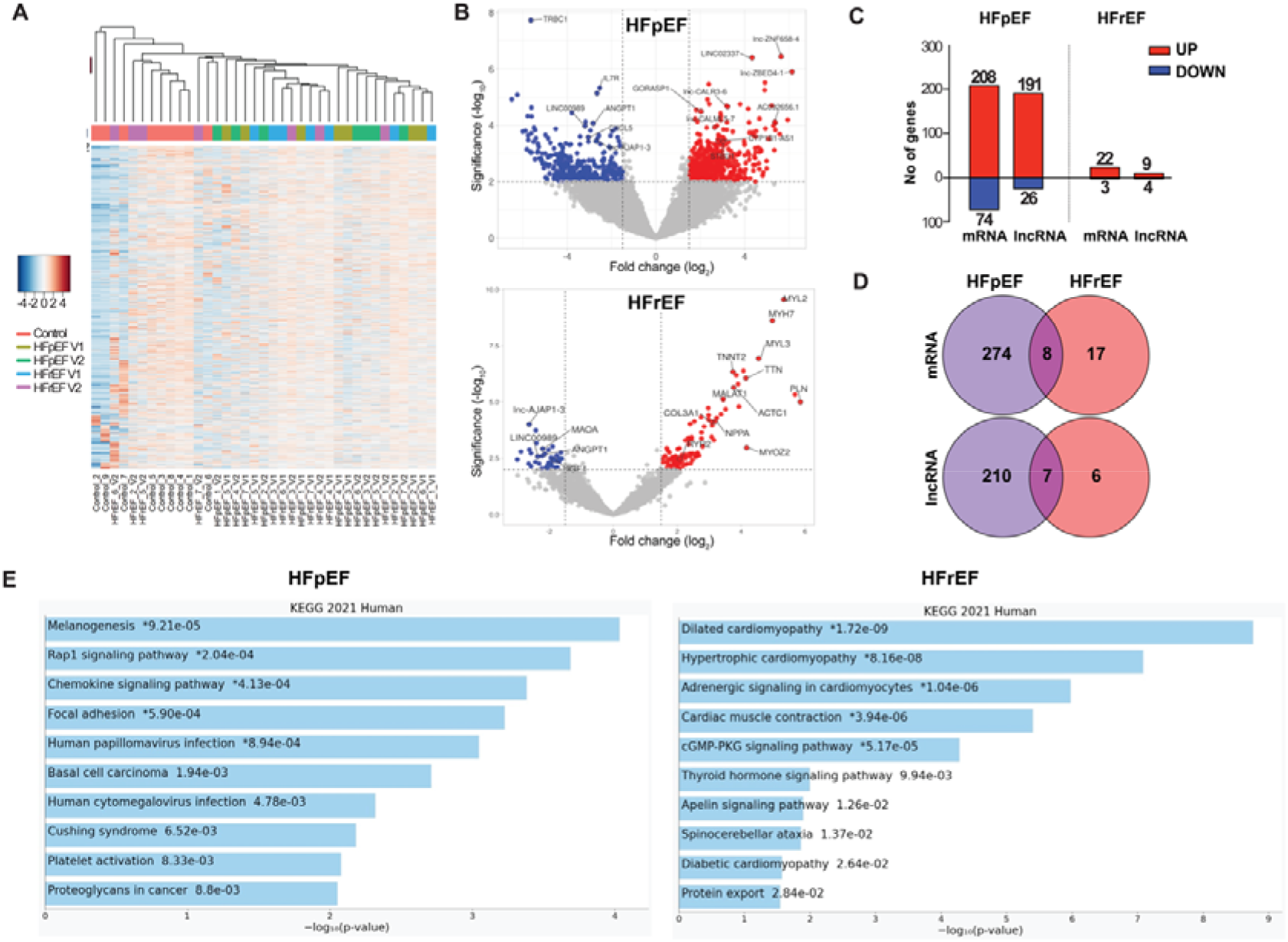
EV long RNA cargo in patients with and without HF (discovery cohort). **A**. Unsupervised hierarchical clustering of all long non-coding RNAs (lncRNAs) in the discovery cohort (Ctrl=9, HFpEF=7, HFrEF=7). V1 and V2 indicate admission (during decompensation) and discharge (after therapy), respectively. **B**. Volcano plots showing the significantly differentially expressed genes from the long RNA sequencing analysis in HFpEF versus Control and HFrEF versus Control indicated as HFpEF and HFrEF, respectively. **C**. Barplot showing up- and down-regulated genes in HFpEF versus Control and HFrEF versus Control indicated as HFpEF and HFrEF respectively. **D**. Commonly expressed mRNAs and lncRNAs between the two analyses (HFpEF versus Control and HFrEF versus Control indicated as HFpEF and HFrEF respectively). **E**. KEGG Pathway enrichment analysis for HFrEF vs Ctrl and HFpEF vs Ctrl. ExRNA indicates Extracellular RNA; Ctrl, Control; HFpEF, Heart Failure with preserved Ejection Fraction; HFrEF, Heart Failure with reduced Ejection Fraction; V1, at Admission (decompensation); V2, at Discharge (after therapy); mRNA, messenger RNA; lncRNA, long non-coding RNA.

### EVs as a source for the differential ADHF transcriptome

Given the ephemeral lifetime of “unprotected” long RNAs in circulation, we hypothesized that circulating long transcripts were mostly associated with EVs or other RNA carriers. We had previously demonstrated the association of small RNAs with different sub-carriers in plasma,^18^ but the association of long RNA transcripts or their fragments had not been examined. Therefore, we isolated RNA from different plasma compartments known to stabilize small RNAs in circulation, including argonaut-2 (Ago2)-associated, lipoprotein-associated, and EV compartments in pooled human plasma from both heart failure and control samples (**Supplementary Figure 1** demonstrates markers associated with each compartment). We validated the EV compartment using MISEV guidelines (in **Figure 3A-C**).^22^ The Minimal Information for Studies of Extracellular Vesicles (MISEV) guidelines prescribe important criteria that need to be fulfilled to validate EV in a study. First, is to characterize the size, and second is to determine the concentration. Both of these were performed using two methods – Transmission Electron Microscopy (TEM, **Figure 3A)**, and Microfluidic Resistive Pulse Sensing (MRPS, **Figure 3B)**.^21^ EVs were isolated using Exoeasy kit and Size Exclusion Chromatography (SEC), measured between 60-100nm in size with a concentration of around 10^10^ EV particles/mL of plasma, although the EVs isolated by the SEC method appeared to be smaller in size. The third MISEV criterion requires the characterization of the EV protein content using western blot with EV markers, specifically, the tetraspanins present on EV-surface (CD81, CD63 or CD9) and EV cargo proteins (Alix, Tsg101 or Syntenin) and, more importantly, for small EVs, a negative marker that includes a soluble intracellular compartment (such as 58KDa Golgi protein). Therefore, we validated our EVs demonstrating the presence of CD81, CD63, Alix, Syntenin, and the absence of 58KDa Golgi protein (**Figure 3C)**. We then probed for the differentially expressed transcripts between HF and control using digital PCR in the different compartments and identified their expression to be enriched in the EV compartment (**Figure 3D**). Finally, the EV markers (examined using western blot) and EV concentrations (examined using Exoview and MRPS methods) were also assessed in our patient groups. This analysis did not reveal any significant differences between the control, HFpEF, and HFrEF groups (**Supplementary Figure 4, 5**) in the baseline characteristics of EVs. Notably, these groups did not differ in total EV concentration, as well as CD81-, CD9- and CD63-positive EVs. Taken together, the data suggest that the plasma concentration of the overall (total) EV populations is not significantly altered in HF patients and that the long RNA transcripts are primarily present in EVs. Since the exRNA transcripts are primarily enriched in EVs, we will refer to them as EV-RNAs henceforth.

**Figure 3.**
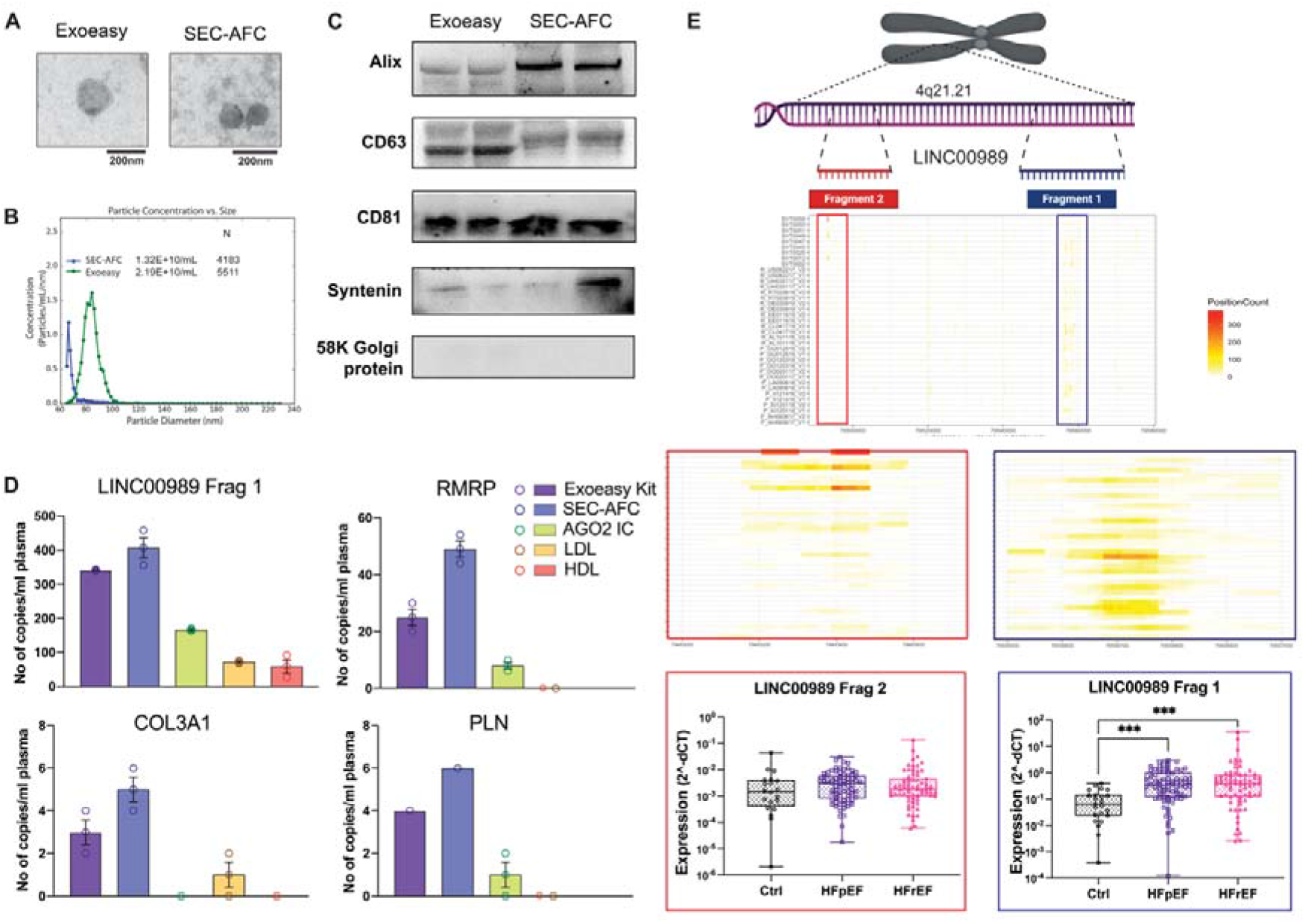
Differentially expressed lncRNA cargo of Extracellular vesicles are often distinctively fragmented. **A**. EV visualization using Transmission Electron Microscope imaging, **B**. EV quantification using MRPS analysis, and **C**. Western blot for EV markers such as Alix, CD63, CD81, and Syntenin as well as “negative” marker 58k Golgi protein reflecting the degree of EV preparation purity. **D**. Analysis of differentially expressed targets from the RNA-sequencing analysis examined by digital PCR in various RNA compartments of pooled human plasma from both heart failure and control samples, namely, EV compartments composed of Exoeasy kit and SEC-AFC; Ago2 -associated RNA compartment obtained by immunocapture using Ago2 antibody; Lipoprotein-associated, LDL and HDL RNA compartments obtained through ultracentrifugation. Validation of the Extracellular Vesicle (EV) compartment using SEC-AFC. **E**. Two lncRNA fragments of the same lncRNA LINC00989 show differential expression patterns in heart failure patient plasma vs. control both in discovery cohort as seen in the coverage plots as well as in the validation cohort evaluated using RT-qPCR. The X-axis shows the full length of the transcript. Y axis represents sample expression. Box plots are obtained using RT-qPCR demonstrated as 2^-dCt values. dCT values are calculated using a Spike-in normalizer. Significance is indicated by * p<0.01 ** p<0.001 ***p<0.0001, where p is calculated using the Kruskal-Wallis test. lncRNAs indicates long non-coding RNA, EV, Extracellular vesicles; RT-qPCR, Real-Time quantitative PCR; Frag 1, Fragment 1; Frag 2, Fragment 2; Ctrl, Control; HFpEF, Heart Failure with preserved Ejection Fraction; HFrEF, Heart Failure with reduced Ejection Fraction; SEC-AFC, Size Exclusion Chromatography using Automated Fraction Collector; AGO2 IC, Immunocapture using Ago2 antibody; LDL, Low Density Lipoprotein; HDL, High Density Lipoprotein; MRPS, microfluidic resistive pulse sensing.

To determine the state of the RNA transcripts within the EVs, we analyzed the expression pattern of the differentially expressed EV-derived RNAs using coverage plots from RNA-sequencing data (**Supplementary Methods**). We would expect uniform coverage of the RNA sequencing reads across the length of the transcript if the full-length transcript were present. Instead, we observed the enrichment of selective fragments suggesting that RNAs were present as fragments within EVs, and that this fragmentation is non-random. For example, while there are two major fragments of LINC00989 detected in our RNAseq data, when we examined the expression of these fragments using primers designed against each of these fragments, only one fragment was differentially expressed between control and HF plasma (**LINC00989 in Figure 3E, Supplementary Figure 6**). While the biological implication of this finding is unclear, it highlights the importance of post-transcriptional processing in RNA biomarker quantification and the importance of using initial RNAseq data to determine the key fragments to analyze using methodologies such as PCR.

### Single nuclear RNA sequencing reveals a cardiomyocyte predominance for top differentially expressed transcripts in ADHF in HFrEF but not HFpEF

Given previous work suggesting HFpEF as a “multi-organ” disease, ^23^ and this broad difference in transcriptional architecture between HF subtypes, we next annotated tissue expression of differentially expressed transcripts. While most differentially expressed transcripts from patients with HFrEF were abundantly expressed in heart and skeletal muscle, transcripts from patients with HFpEF exhibited a broader variety in expression across divergent tissue types (**Figure 4A-D**), consistent with clinical observations of its rich pathophysiologic heterogeneity. We next “deconvoluted” differentially expressed transcripts using single nuclear RNA quantification in the human heart (non-failing, DCM, and HCM).^19^ DCM is characterized by dilation and contractile dysfunction of one or both ventricles, resulting in HFrEF, while HF manifestations of HCM primarily phenocopy HFpEF. Most of the top differentially expressed transcripts from HFrEF patients were of cardiomyocyte origin, primarily composed of several genes involved in sarcomeric function (ACTC1, ACTN2, MYL2, MYL3, MYL7, MYH6, MYH7, PLN, TNNC1, TNNT2, TTN) (**Figures 4E-H;** UMAP legend in **Figure 4I**). By comparison (and consistent with our bulk tissue of origin data in **Figure 4D**), most differentially expressed RNAs from patients with HFpEF were expressed in a broad array of non-cardiomyocyte cell types within the heart, including endothelial cells, fibroblasts (ANGPT1), and immune cells (CD226, IL7R) (**Figures 4F, 4H)**. While the current methodology of single nuclear RNAseq does not provide transcript data on RNAs without polyA tails, we were able to assess the possible cellular origin of a few of our differentially expressed lncRNAs. For instance, LINC02337 was detected ubiquitously in all the different types of cardiac cells. In contrast, LINC00989, expressed by fewer cardiomyocytes, was predominantly expressed by the cardiac pericytes. RMRP was detected in many cell types of cardiac cells with abundant expression in cardiomyocytes. ST20-AS1, is expressed much less in various cell types, with expression primarily in cardiomyocytes and fibroblasts.

**Figure 4.**
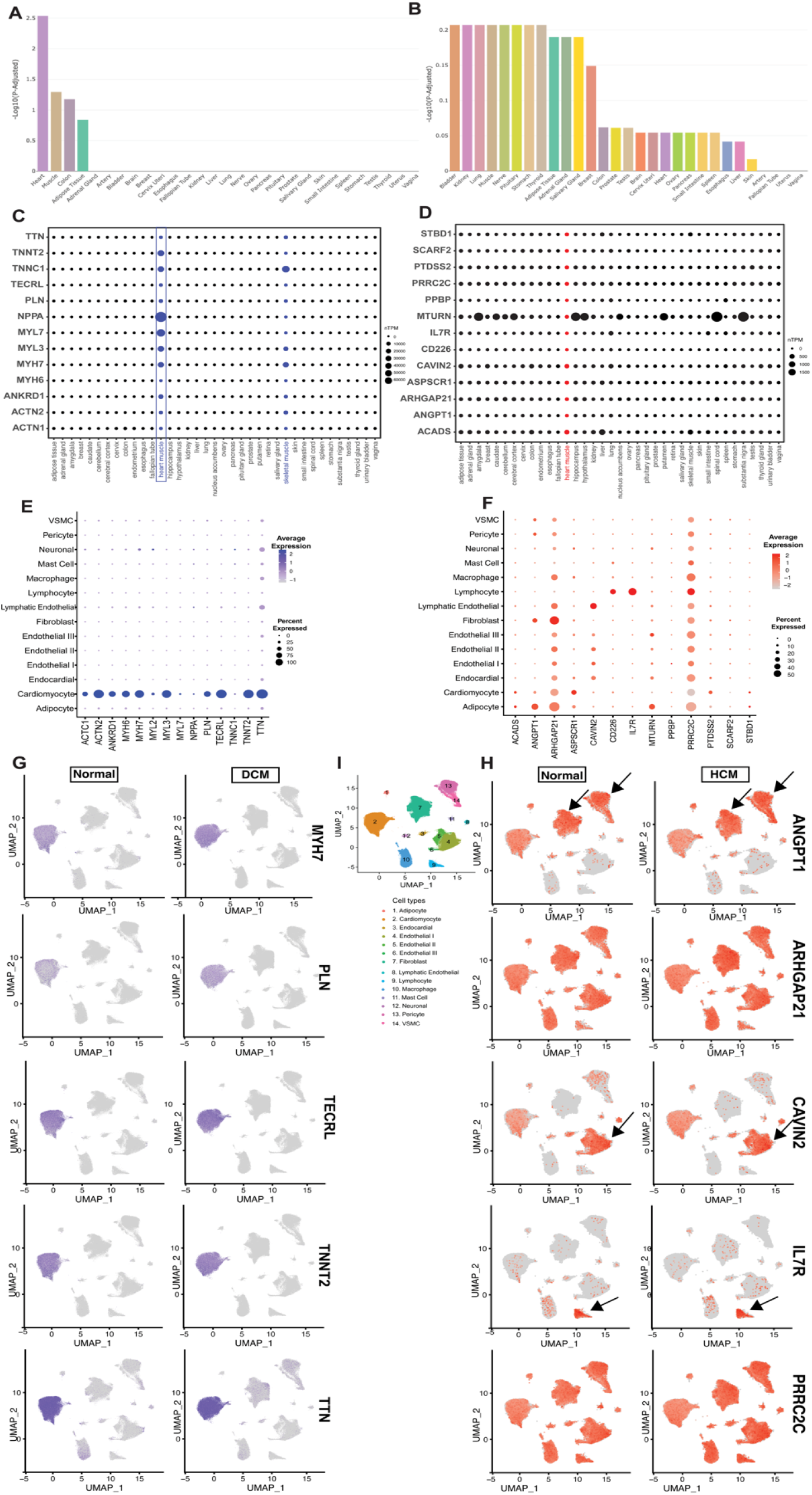
Enrichment analysis of DEGs in plasma-derived EVs of heart failure subtypes deconvoluted using single nuclear profiling of human hearts. Bar plots showing the tissue enrichment analysis data (from GTEx tissue RNA atlas) of the topmost upregulated transcripts between **A**. HFrEF EVs vs Ctrl EVs and **B**. HFpEF EVs vs Ctrl EVs (obtained from TissueEnrich). Dot plot analysis showing tissue enrichment data (from GTEx tissue RNA atlas) of the topmost differentially expressed genes from **C**. HFrEF EVs vs Ctrl EVs and **D**. HFpEF EVs vs Ctrl EVs. Dot plot analysis from the single nuclear transcriptomic analysis of the topmost differentially expressed genes from **E**. HFpEF EVs vs Ctrl EVs and **F**. HFrEF EVs vs Ctrl EVs. **G**. Representative Uniform manifold approximation and projection plots (UMAP) of top DEG in HFrEF vs Ctrl analysis as observed in normal and DCM hearts; and **H**. Representative UMAPs of top DEG in HFpEF vs Ctrl analysis as observed in normal and HCM hearts. **I**. UMAP legend: heart tissue cellular map composed of 14 different types of cardiac cells. EVs indicate Extracellular vesicles; Ctrl, Control; HFpEF, Heart Failure with preserved Ejection Fraction; HFrEF, Heart Failure with reduced Ejection Fraction; DCM, Dilated Cardiomyopathy; HCM, Hypertrophic Cardiomyopathy.

### Validation of differentially expressed RNAs in HF subtypes

Next, we measured the top differentially expressed RNAs using qRT-PCR on plasma-derived EVs obtained at admission from our validation cohort. Expression of 13 targets was examined in 182 patients (24 control; 86 HFpEF; 72 HFrEF), out of which 11 targets (5 lncRNAs and 6 mRNAs) were significantly different between control and ADHF after adjustment for age and sex, stratified by HF subtype (**Figure 5A, 5B**). To enhance the rigor of the results obtained, we further used digital PCR to confirm the qRT-PCR results (**Supplementary Figure 7)**. Interestingly, these EV-RNAs were altered in a similar direction in HFpEF and HFrEF, and receiver-operating characteristic (ROC) analysis performed on the top five validated targets for each of the HF subtypes after adjustment for age and sex (0.9301 for HFpEF and 0.8617 for HFrEF) improved the C-statistics (**Figure 5C**) when compared to age and sex alone (0.8176 for HFpEF and 0.6369 for HFrEF) (**Supplementary Figure 8**).

**Figure 5.**
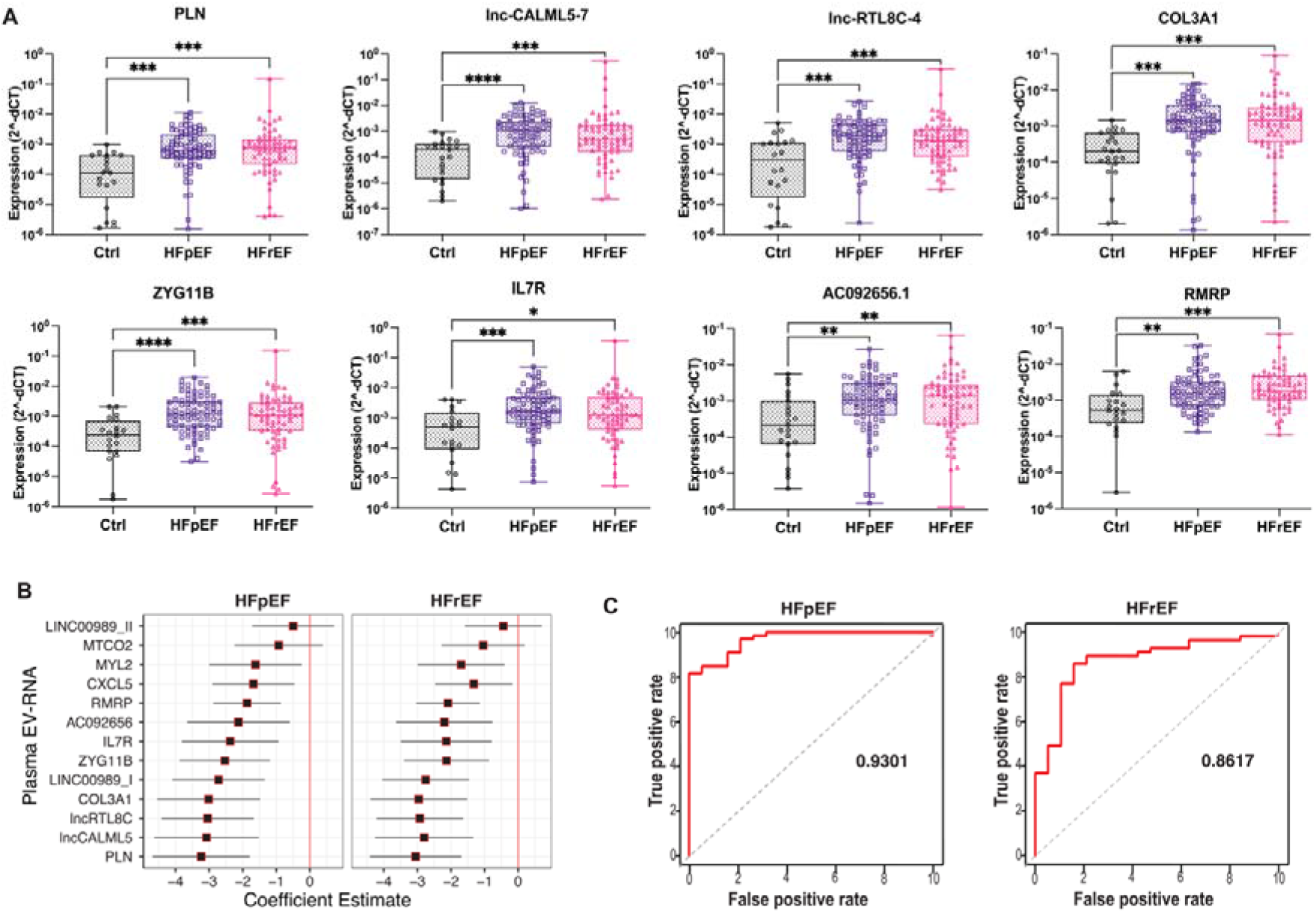
EV-derived transcripts were differentially expressed between control and acute decompensated heart failure patients in the Validation cohort. **A**. Box plots showing some of the differentially expressed targets that were experimentally confirmed on the validation cohort (Ctrl=24, HFpEF=86, HFrEF=72) using RT-qPCR demonstrated as 2^-dCt values. dCT values are calculated using a Spike-in normalizer. Significance is indicated by * p<0.01 ** p<0.001 ***p<0.0001, where p is calculated using the Kruskal-Wallis test. **B**. Forest plot of all the experimentally validated differentially expressed EV cargo. The coefficient estimate represents the numerical differences between each HF subtype and control adjusted for age and sex. Negative values represent decreasing dCT values or increased plasma EV-RNA expression. **C**. Diagnostic ROC curves for the five topmost validated targets between Control and Heart Failure Subtypes at Acute decompensated state. EV indicates Extracellular vesicles; RT-qPCR, Real-Time quantitative PCR; I, Fragment 1; II, Fragment 2; Ctrl, Control; HFpEF, Heart Failure with preserved Ejection Fraction; HFrEF, Heart Failure with reduced Ejection Fraction; dCT, delta CT value.

### Plasma EV lncRNAs (but not mRNAs) exhibit dynamic expression during decongestion

To understand whether the transcripts that validated during the admission of the HF subtypes were a marker of congestion status or reflected other pathophysiological markers during ADHF, we examined changes in expression during decongestion. We prioritized transcripts that were most differentially expressed in HF patients at the congested (acute) state and analyzed their expression (via qRT-PCR) after decongestion (diuretic treatment). Of the 11 targets examined, only 4 non-coding RNAs (LINC00989, lnc-CALML5, AC092656.1, RMRP) changed during therapy for ADHF (**Figure 6A, Supplementary Figure 9**). The expression of LINC00989 and RMRP decreased with therapy, and the expression of lnc-CALML5 and AC092656.1 increased. Interestingly, while there was a significant decrease in body weight with decongestion in both HFpEF and HFrEF patients with ADHF, NT-proBNP was significantly decreased only in the HFrEF patients (**Supplementary Figure 10)**, confirming the challenges associated with NT-pro-BNP use in HFpEF.^24^ EV-derived lncRNAs were only weakly correlated with weight change or change in NT-proBNP (data not shown).

**Figure 6.**
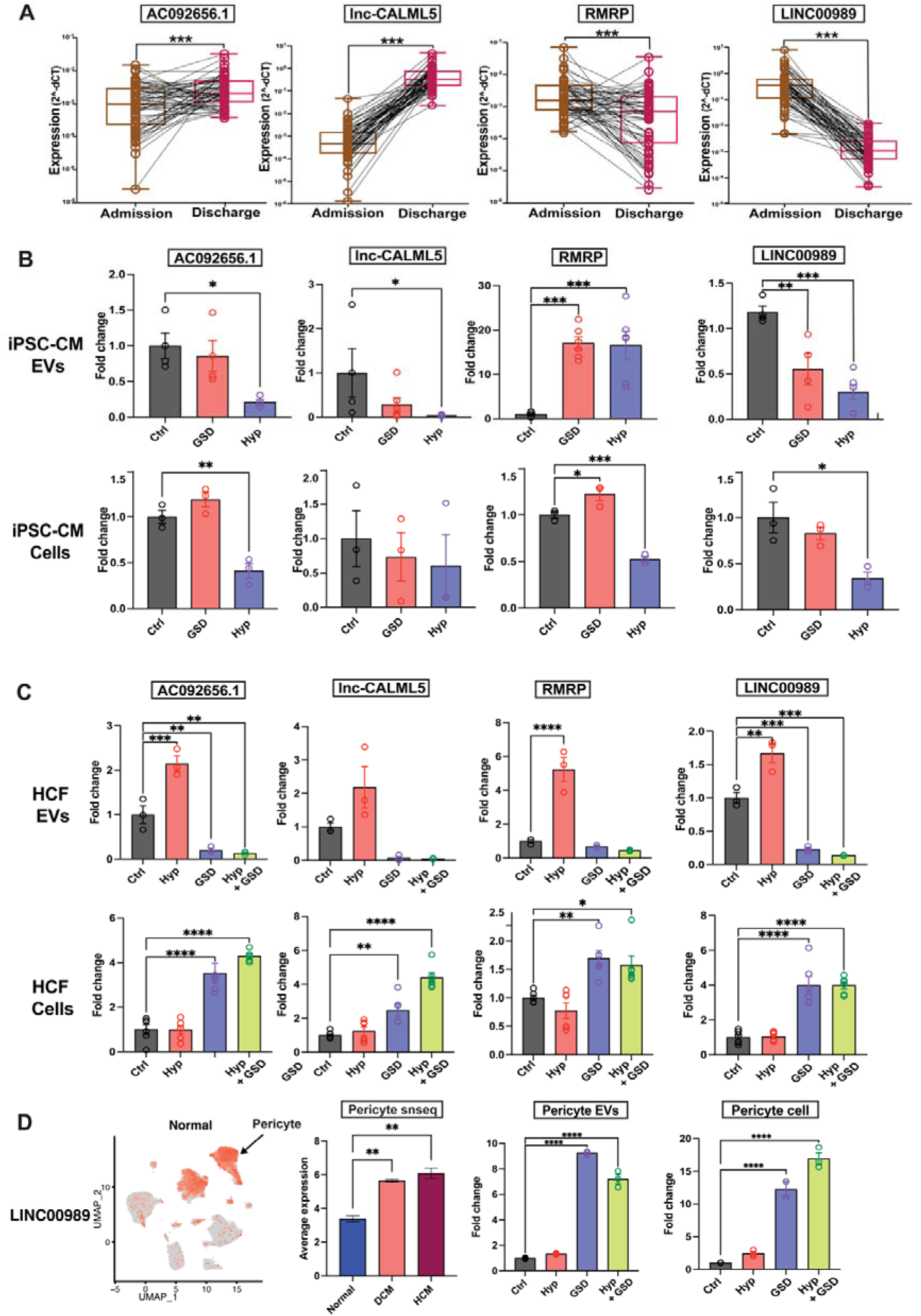
EV-derived lncRNAs or their fragments change dynamically and significantly between acute congested to decongested states. **A**. Box plots of the differentially expressed targets that were experimentally confirmed on the validation cohort between admission and discharge using RT-qPCR. Significance expressed as * p<0.01 ** p<0.001 ***p<0.0001 where p is calculated using the Kruskal-Wallis test. EV and cellular RNA expression of AC092656.1, lnc-CALML5-7, RMRP, and LINC00989 upon hypoxia and nutrient deprivation (Glucose serum deprivation) in **B**. Induced Pluripotent Stem Cell-derived cardiomyocytes (iPSC-CMs) and **C**. Human Cardiac Fibroblasts (HCF) assessed using qRT-PCR. Fold change is calculated using the 2^−ΔΔCt^ method relative to 5s rRNA in EVs and BACT in cells. Significance is expressed as * p<0.01 ** p<0.001 ***p<0.0001 where p is calculated using ANOVA test. **D**. UMAP plot of LINC00989 from single nuclear transcriptomic data showing expression in pericytes; Expression of LINC00989 in normal, DCM, and HCM pericytes from single nuclear transcriptomic data; Expression of LINC00989 in pericyte EVs and pericyte cells subjected to stressors obtained from qRT-PCR as mentioned above. EV indicates Extracellular vesicles; RT-qPCR, Real-Time quantitative PCR; Ctrl, Control; HFpEF, Heart Failure with preserved Ejection Fraction; HFrEF, Heart Failure with reduced Ejection Fraction; HF, Heart Failure; dCT, delta CT values; Hyp, Hypoxia; GSD, Glucose Serum Deprivation.

### Dynamic changes in LINC00989, RMRP, lnc-CALML5-7 and AC092656.1 with cellular stress in human induced pluripotent stem cell-derived cardiomyocyte, human cardiac fibroblast, and human pericyte models

Given that these four lncRNAs dynamically change from the acute state to decongested states, we hypothesized that expression of these lncRNAs could be altered at the cellular level when subjected to different stress conditions such as hypoxia (Hyp) and nutrient deprivation (glucose serum deprivation or GSD). Upon subjecting induced pluripotent stem cells-derived cardiomyocytes (iPSC-CMs) to the cellular stressors, we found that the expression of lnc-CALML5-7, AC092656.1, and LINC00989 in EVs and in cells was decreased under hypoxic stress (coordinate with the changes in ADHF for the first two, where plasma EV levels showed lower expression of lnc-CALML5-7 and AC092656.1). In contrast, the expression of RMRP increased in iPSC-CM EVs during stress, similar to the higher levels in plasma EVs of patients with ADHF, while its cellular levels showed a decrease (**Figure 6B**). We also examined the cellular and EV expression of these lncRNAs in other cardiac cell-types. When human cardiac fibroblasts (HCF) were subjected to cellular stressors, the expression of all the four lncRNAs in EVs increased upon hypoxia and decreased upon GSD while the converse trend was observed at the cellular level where their expression increased during GSD (**Figure6C)**. Since acute decompensation is a form of acute stress for patients with HF, the trends observed in stressed iPSC-CM EVs for AC09265.18, lnc-CALML5-7, and RMRP are concordant with those observed in the plasma of HF patients but not for LINC00989. Interestingly, the single nuclear transcriptomic analysis revealed that LINC00989 was highly expressed in cardiac pericytes and not in cardiomyocytes. We, therefore, also examined the expression of LINC00989 in these cells. When pericytes were subjected to cellular stressors, the expression of LINC00989 was elevated in both cellular and EV compartments. This is in accordance with the increased expression of LINC00989 in patient plasma during the acute congested state (**Figure 6D)**.

## DISCUSSION

Patients with HF experience poorer survival after an acute decompensation,^1^ though whether these findings represent inadequate decongestion or systemic-myocardial signaling that contributes to a declining cardiac function remains unclear. Despite their discovery more than five decades ago, a functional role for EVs in HF is new.^25,26^ There are two broad principal findings from our study. First, we observed that the HFpEF and HFrEF EV transcriptome is distinct and exhibits organ- and cell-type specificity in the acutely congested state that reflects a divergent underlying pathophysiology. Second, EV lncRNA transcripts change with decongestion, and are altered with stress in appropriate human-derived cells (iPSC-derived cardiomyocytes, and pericytes) in a direction similar to that observed in patients during the acute-to-recompensated transition. While deconvolution approaches have been used in other contexts to define EV origins,^27^ its use here as a “liquid biopsy” across HF subtypes suggested that while the EV transcripts in decompensated HFrEF were more clearly expressed in the heart, specifically in cardiomyocytes, those in decompensated HFpEF exhibited broader, systemic origins (non-cardiomyocyte) - in line with current clinical conception of HFpEF as a multi-systems disorder. Furthermore, the transcripts prioritized based on patient-oriented EV findings during decongestion demonstrated dynamic *in vitro* expression, which further underscores the possibility that select lncRNA expression profiles may be specifically implicated in myocardial responses to stress in HF. Collectively, these results demonstrate dynamic transitions in the EV transcriptome across acute to decongested states that reflect a potential divergent HF pathophysiology (“liquid biopsy”), with cellular model evidence that prioritizing EV transcriptome changes during HF therapy may inform discovery efforts across HF sub-types.

Most efforts to define the transcriptional architecture of HF have focused on the myocardium,^6,7,19,28^ with the hypothesis that changes within the myocardial tissue may identify intervenable pathways of cardiac recovery and therapeutic response. While these seminal efforts have undoubtedly led to dramatic advances in understanding diverse patterns and mechanisms of genetic regulation in the heart, they are generally cross-sectional in end-stage cardiomyopathy or are limited by potential confounders such as cross-sample differences in HF risk that may contribute to altered expression independent of the HF state (e.g., BMI). In response, circulating “omic” biomarkers of HF—both acute and chronic— have been proposed to resolve heterogeneities in risk.^23^ While these biomarkers may be accessible for serial sampling and more easily translatable to the clinic, both metabolomics and proteomics may not have sufficient mechanistic or tissue specificity to prioritize targeting efforts. In addition, recent large studies suggest a lack of association between human genetic variation and subtypes of HF (HFpEF vs. HFrEF), suggesting that more phenotype-adjacent marks of HF status may provide more resolution.^5^ The <u>circulating</u> transcriptome has therefore been an area of active mechanistic investigation in human HF. Previous studies utilizing cell-based transcriptomics (e.g., peripheral immune profile) or “cell-free” transcriptomics (e.g., circulating miRNAs, lncRNAs) have been successful (specifically with lncRNAs^29^), though with variable results likely due to several factors such as single time-point study, low-input quantity, RNA fragmentation, and differences in quantification platform and population. Importantly, these few studies have used PCR-based array data to identify known lncRNAs in the circulation that may be associated with chronic HF: the mitochondria-derived lncRNA LIPCAR was found to be associated with left ventricular remodeling after myocardial infarction^29^ and circulating long non-coding RNAs NRON and MHRT were identified as predictors of HF.^30^ To our knowledge, our study is the first to employ RNAseq to identify EV transcripts as an unbiased approach to discovery. Finally, our approach addresses a key confounder in plasma transcriptome studies: given that fragments rather than full-length RNAs are present in the EV compartment, PCR based approaches for circulating transcript detection may be insensitive or lack rigor without detailed prior analysis of the exact fragments (coverage) present in the compartment.

By analogy to oncologic approaches with EVs,^31,32^ we hypothesized that the EV compartment would serve as a “liquid biopsy” that is a barometer of the HF state. Plasma transcripts enriched in HFrEF patients were primarily present in the EV compartment and appeared cardiac-specific (predominantly in cardiomyocytes), whereas those in HFpEF were also present in EVs, but were of diverse cellular origin. In addition, the HFrEF transcriptome appeared functionally distinct from that of HFpEF—converging on heart muscle-enriched pathways—while the HFpEF transcriptome pointed to a broader pathway array (e.g., Rap1 signaling, chemokine signaling, etc.). Of note, these findings mirror observations from myocardial biopsy, with more cardio-specific pathways altered in HFrEF (e.g., dysregulated cytoskeletal structural heart genes^33^) and a wider mechanistic berth encompassing several non-structural pathways associated with mitochondrial ATP synthesis, electron transport pathways, endoplasmic reticulum stress, and autophagy in HFpEF.^34^

Moreover, despite these clear differences between HFpEF and HFrEF, we observed a convergence in how the EV transcriptome responds to HF therapy across HF subtypes for the top lncRNA targets. We found that expression of these top lncRNAs—LINC00989, RMRP (both decreased); lnc-CALML5-7, AC092656.1 (both increased)—were dynamically expressed in EVs between a congested and decongested state in both HF subtypes. While the basis for this similarity is speculative, given the importance of the ADHF moment in the prognosis of HF across both subtypes, it is tantalizing to hypothesize a role for these RNAs in cellular processes common between the two subtypes (such as fibrosis or inflammation), or alternatively a role for EV RNAs in RNA quality control.

While lncRNAs represent a large fraction of the altered myocardial transcriptome in HF,^7^ there are few studies cataloging their mechanistic ontology, given poor murine to human homology. Human RMRP is 264 bp-sized lncRNA with nuclear or mitochondrial localization involved in miRNA generation and transcriptional and genomic processing.^35–37^ In HF, RMRP has been implicated in hypertrophy,^35,38^ fibrosis,^39^ and ischemia. ^40^ While studies on the murine homolog suggest conflicting, context-dependent roles,^39,41^ prior work in human demonstrates an increase in RMRP after ischemia. ^40^ We observed primarily cardiomyocyte expression in snRNA-seq, and dynamic regulation in human *in vitro* models under metabolic stress. LINC00989 is primarily expressed in cardiac pericytes (from snRNA-seq), though its role in HF is unknown. However, cardiac pericytes have been implicated in myofibroblast activation^42,43^ and epithelial-mesenchymal transition (EMT)^44^ across various diseases.^45,46^ Also, *in vitro* experiments using our patient-oriented studies suggested dynamic expression of LINC00989 with stress in human pericytes, potentially implicating lncRNA in cardiac pericyte-mediated roles in HF.^20,21^ While the other RNAs identified remain functionally occult: lncCALML5-7 is an intergenic lncRNA proximal to CALML3, a calmodulin-like protein 3, and AC092656.1 is a pseudogene of Mitochondrial Ribosomal Protein L42 (MRPL42). Interestingly, CALML3 protein is expressed at low levels in heart and has been reported to bind to KCNQ1 to form functional octamers to act as Ca^2+^ sensor in epithelial cells. However, cis-dependent association of CALML3 with lnc-CALML5-7 in the context of cardiomyocytes has not been previously assessed. Finally, AC092656.1 has an interesting association with MYOZ2, where the lncRNA is found on the reverse strand, while the protein counterpart is found on the forward strand. MYOZ2 has been associated with cardiomyopathy. However, this is the first study to correlate AC092656.1 expression with heart failure. Given the broad diversity of lncRNAs in tissue and circulation, disparate post-transcriptional regulatory functions of lncRNAs, and difficulty in their study (species-specificity) necessitating human studies, the approach laid out in this work—plasma, tissue, and EV compartment—offers a roadmap on prioritization of targets for further study from human circulation to potential human biology.

Several important limitations merit mention. We have used two different methodologies with detailed characterization of the EV isolates to support our claim that unlike other small RNAs, mRNA/lncRNA transcripts are present mostly in EVs; nonetheless, given inherent difficulties in the field of isolating ‘pure’ EVs without contaminating co-isolates, we concede that these RNA fragments may also be present to a smaller extent in association with other RNA carriers. While our study may be viewed as descriptive, observing a distinct architecture between HFpEF and HFrEF through deconvolution of EV transcriptional profiles that identify genes responsive to stimuli in human cardiomyocyte cell systems is novel and consistent with clinical observation. Given limitations in snRNA-seq data available for deconvolution, we used hypertrophic cardiomyopathy snRNA sequencing for deconvolution of myocardial cell specificity representative of HFpEF; nonetheless, HFpEF EV transcripts did not exhibit an enrichment in the heart in general samples (e.g., GTEx, Figure **4A-D**). As with other studies,^6^ patient heterogeneity in HFpEF (eg., BMI, age) and therapy are potential confounders, which highlights a strength of our study—serial, within-individual testing of how EV transcripts change with therapy, which may be more robust to these differences (a study design not ethically possible with heart biopsy). We attempted to adjust for these conditions in our modeling. The sample size in our discovery group was small, but the study was longitudinal with replicated effects in validation cohort and dynamic alterations *in vitro*.

In conclusion, the circulating EV transcriptome is significantly altered during acute HF, with a distinct cell and organ specificity in HFpEF vs. HFrEF consistent with a multi-organ vs. cardiac origin, respectively. Plasma EV-derived lncRNA fragments were more dynamic with acute HF therapy independent of weight change (relative to mRNAs) and demonstrated dynamicity with cellular stress *in vitro*. Prioritizing transcriptional changes in plasma circulating EV with HF therapy may be a fruitful approach to HF subtype-specific mechanistic discovery.

## Data Availability

All data produced in the present study are available upon reasonable request to the authors

## ACKNOWLEDGEMENTS

We acknowledge the contribution of MGH Core facilities for sequencing and microscopy services.

## Graphical abstract

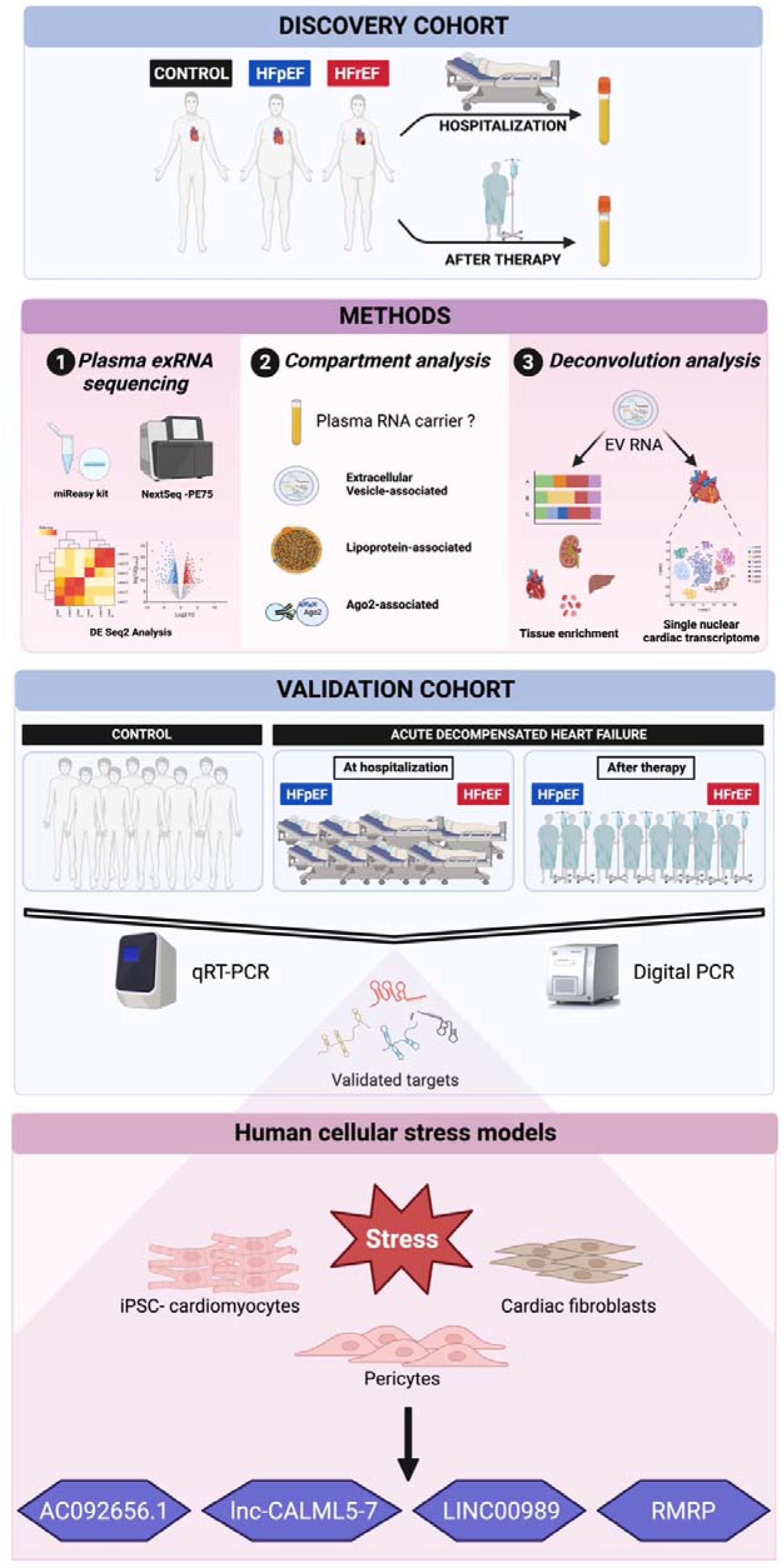

## Supplemental Material

Supplemental Methods

Supplementary Tables S1–S3

Supplementary Figure S1-S10

References 46-50

